# Obesity and Smoking as Risk Factors for Invasive Mechanical Ventilation in COVID-19 Respiratory Failure: a Retrospective, Observational Cohort Study

**DOI:** 10.1101/2020.08.12.20173849

**Authors:** Ana C. Monteiro, Rajat Suri, Iheanacho O. Emeruwa, Robert J. Stretch, Roxana Y. Cortes-Lopez, Alexander Sherman, Catherine C. Lindsay, Jennifer A. Fulcher, David Goodman-Meza, Anil Sapru, Russell G. Buhr, Steven Chang, Tisha Wang, Nida Qadir

## Abstract

**Purpose:** To describe the trajectory of respiratory failure in COVID-19 and explore factors associated with risk of invasive mechanical ventilation (IMV).

**Materials and Methods:** A retrospective, observational cohort study of 112 inpatient adults diagnosed with COVID-19 between March 12 and April 16, 2020. Data were manually extracted from electronic medical records. Multivariable and Univariable regression were used to evaluate association between baseline characteristics, initial serum markers and the outcome of IMV.

**Results:** Our cohort had median age of 61 (IQR 45-74) and was 66% male. In-hospital mortality was 6% (7/112). ICU mortality was 12.8% (6/47), and 18% (5/28) for those requiring IMV. Obesity (OR 5.82, CI 1.74-19.48), former (OR 8.06, CI 1.51-43.06) and current smoking status (OR 10.33, CI 1.43-74.67) were associated with IMV after adjusting for age, sex, and high prevalence comorbidities by multivariable analysis. Initial absolute lymphocyte count (OR 0.33, CI 0.11-0.96), procalcitonin (OR 1.27, CI 1.02-1.57), IL-6 (OR 1.17, CI 1.03-1.33), ferritin (OR 1.05, CI 1.005-1.11), LDH (OR 1.57, 95% CI 1.13-2.17) and CRP (OR 1.13, CI 1.06-1.21), were associated with IMV by univariate analysis.

**Conclusions:** Obesity, smoking history, and elevated inflammatory markers were associated with increased need for IMV in patients with COVID-19.

## Introduction

Coronavirus disease 2019 (COVID-19) has been reported in over 200 countries,^1^ leading to an unprecedented impact on healthcare systems worldwide. While its disease course is incompletely understood, its causative virus, SARS-CoV-2, is thought to enter respiratory epithelial cells via the angiotensin-converting enzyme-2 (ACE2) receptor in the lungs, resulting in a variety of clinical symptoms. Given the high infection rates worldwide, the identification of markers predicting increased disease severity may enable more effective triaging of patients and procurement of resources.

Due to its novelty and heterogeneity in presentation, the underlying mechanism for the severe hypoxic and hypercapnic respiratory failure sometimes seen in COVID-19 has been the subject of controversy. The common features of bilateral pulmonary infiltrates in combination with low PaO2/FiO2 (P/F) ratios are consistent with acute respiratory distress syndrome (ARDS). However, other potential etiologies for COVID-19-related respiratory failure have been proposed, including an atypical, “high compliance” form of ARDS, high-altitude pulmonary edema, and other non-pulmonary causes of hypoxia, such as those caused by cardiomyopathy.^2-5^ The presence of various hypotheses for disease mechanism may be playing a role in the management and outcomes of patients with respiratory failure.

We present our cohort of the first 112 unique, consecutively admitted patients to our hospital system with confirmed COVID-19 infection. Our aim was to describe characteristics, management, and trajectory of respiratory failure and mortality in our cohort, and to explore the factors associated with the need for invasive mechanical ventilation.

## Methods

### Setting, Patient Population, and Study Design

This is a retrospective observational cohort study conducted from March 12 2020 and April 16 2020 approved by the UCLA (University of California, Los Angeles) institutional review board with waiver of informed consent. The UCLA hospital system is an academic center in Los Angeles County comprised of Ronald Reagan-UCLA Medical Center (RR-UCLA) and Santa Monica-UCLA Medical Center (SM-UCLA). RR-UCLA has 520 beds, of which 109 are critical care beds, while SM-UCLA has 281 beds, of which 22 are ICU beds. Hospitalized patients at RR-UCLA and SM-UCLA ≥ 18 years old with positive SARS-CoV-2 PCR testing from either nasal swab or mini-bronchoalveolar lavage (BAL) testing were included. Data for patients who tested positive for SARS-CoV-2 was manually extracted from the electronic health record and included in a database.

### Clinical Protocols

This was an observational study, and all clinical management was left to the discretion of the primary treatment team. A pandemic response team comprised of intensivists and infectious disease specialists generated COVID-19 treatment guidance documents; recommendations emphasized established best critical care practices. Recommendations for patients who developed acute respiratory distress syndrome (ARDS) included low tidal volume ventilation with tidal volumes ≤ 6 mL/kg predicted body weight (PBW), early consideration of prone ventilation for patients with P/F ratios < 150, daily awakening trials or light sedation, and conservative fluid management. Titration of positive end-expiratory pressure (PEEP) was recommended to reflect ARDS network PEEP tables.^6-9^

All admitted patients with COVID-19 were considered for enrollment in interventional clinical trials. Clinical trials available at our centers from March 12, 2020 to April 16, 2020 included those testing the therapeutic benefit of sarilumab (NCT04315298), remdesivir (NCT 04280705), leronlimab (NCT04347239) and hydroxychloroquine (NCT04332991). In addition, during this time, several patients received open-label remdesivir, leronlimab, hydroxychloroquine and tocilizumab (Supplemental Tables 1 and 2).

### Data Collection

Baseline demographic variables, smoking history, comorbidities, and body mass index (BMI) were collected. Inflammatory markers including C-reactive protein (CRP), D-dimer, ferritin, lactate dehydrogenase (LDH) and highly-sensitive Interleukin-6 (IL-6); and other labs such as hemoglobin A1c, troponin, procalcitonin and a white blood cell count with differential were obtained on admission, or on day of COVID-19 diagnosis if the cause of the original admission was unrelated to confirmed or suspected COVID-19. Treatment-level variables collected included use of anticoagulation, enrollment in clinical trials, as well as the use of open-label directed therapies for COVID-19, which included hydroxychloroquine, tocilizimab, leronlimab, and remdesivir. For those patients on mechanical ventilation, ventilator and respiratory parameters were collected, as were the use of adjunctive ARDS therapies, including prone positioning, neuromuscular blockade, pulmonary vasodilators, and extracorporeal support. Outcomes data that were collected included 60 day in-hospital mortality, ICU admission, rate of endotracheal intubation, ICU length of say, hospital length of stay, and duration of mechanical ventilation. We defined adherence to lung protective ventilation as the use of tidal volume (V_T_) < 8 mL/kg predicted body weight (PBW).

### Definitions

We identified patients who had ARDS as defined by the Berlin Criteria—P/F ratio < 300 with chest imaging revealing bilateral opacities that could not be exclusively explained by cardiogenic causes.^10^ Diagnosis of venous thromboembolism (VTE) was considered present if there was documented radiographic evidence of either pulmonary embolism (via computerized tomography) or deep venous thrombosis (via ultrasound). We defined mortality as death during the first 60 days of inpatient hospitalization. Any admission to the ICU or any intubation for respiratory failure, regardless of duration, was included in the rate of ICU admissions and intubations, respectively.

### Statistical Methods

We combined key categorical variables so that smoking history was evaluated as present, former, or never-smoker, and comorbidities as no known medical history versus any past medical history. We also evaluated these categorical variables in all their individual categories when providing descriptive statistics. Race/ethnicity was defined as White, Black, Latinx, Asian, or other. Descriptive statistics employing simple mean, median and interquartile range were used for the baseline characteristics of the whole cohort and by intubation status. Two-by-two tables were used to describe outcomes of selected subgroups. Chi-squared analysis was used to assess statistical significance between categorical variables, two sample t-test and two sample Wilcoxon Rank-Sum tests were used to assess statistical significance between continuous variables. Wilcoxon Rank-Sum test was used when the total number was low and the assumption of a normal distribution was not met.

Multivariable logistic regression was utilized to assess the contribution of selected baseline characteristics of the cohort to the odds of the binary outcome of requiring mechanical ventilation. Past medical history with cohort prevalence of >15% were used in the multivariable model. We also explored association between selected pro-inflammatory markers and odds of requiring mechanical ventilation by using univariate logistic regression models for each biomarker. Multivariable analysis was not used for biomarkers because of significant co-linearity between the inflammatory markers. We established a threshold of 15% missingness for key variables of interest *a priori* to trigger multiple imputation, which was not met for any variable in our analyses.

## Results

### Patient Characteristics

We evaluated the first 113 unique admissions to our healthcare system with confirmed COVID-19 infection, as diagnosed between March 12, 2020 and April 16, 2020. We excluded one patient who incidentally tested positive for COVID-19 but died from complications from a motor vehicle collision before COVID-directed inpatient management was initiated.

Among the remaining 112 patients, the median age was 61 years old (IQR 45-74) and subjects were predominantly male (66%). The cohort was 44% White, 29% Latinx, 8% Asian, 6% Black, and 13% other. The majority (84%) had a known comorbidity on admission, with the most frequent comorbidities being diabetes (65%), hypertension (50%), obesity (36%), chronic kidney disease (17%), and coronary artery disease (15%) (Table 1). Of our cohort, 27% were taking either ACE-inhibitors or angiotensin receptor blockers (ARBs) as an outpatient, and 7% were health care workers. Four patients were admitted as a transfer from an outside hospital for higher level of care.

**Table 1.** Baseline Characteristics of Cohort.

|  |  | Total (N=112) | Never<br>Intubated<br>(N=84) | Intubated<br>(N=28) | P-value |
| --- | --- | --- | --- | --- | --- |
| **Sex** | Male | 74 (66%) | 55 (65%) | 19 (68%) | 0.818 |
|  | Female | 38 (34%) | 29 (35%) | 9 (32%) |  |
| **Age** |  | 61 (45-74) | 64 (43-77) | 58 (51-64) | 0.320 |
| **Race/Ethnicity** | White | 49 (44%) | 41 (49%) | 8 (29%) | 0.350 |
|  | Latinx | 33 (29%) | 24 (29%) | 9 (32%) |  |
|  | Asian | 9 (8%) | 6 (7%) | 3 (11%) |  |
|  | Black | 7 (6%) | 4 (5%) | 3 (11%) |  |
|  | Other | 14 (13%) | 9 (11%) | 5 (18%) |  |
| **Past Medical History** | No PMHx | 17 (15%) | 14 (17%) | 3 (11%) | 0.447 |
|  | Obesity | 40 (36%) | 23 (27%) | 17 (61%) | 0.001 |
|  | Hypertension | 56 (50%) | 39 (46%) | 17 (61%) | 0.190 |
|  | Diabetes | 72 (64%) | 53 (63%) | 19 (68%) | 0.649 |
|  | COPD | 6 (5.4%) | 4 (5%) | 2 (7%) | 0.628 |
|  | CAD | 17 (15%) | 14 (17%) | 3 (11%) | 0.447 |
|  | Cancer | 15 (13%) | 13 (15%) | 2 (7%) | 0.262 |
|  | Asthma | 13 (12%) | 9 (11%) | 4 (14%) | 0.609 |
|  | Atrial Fibrillation | 11 (10%) | 10 (12%) | 1 (4%) | 0.199 |
|  | CKD | 19 (17%) | 16 (19%) | 3 (11%) | 0.309 |
|  | Transplant Recipient | 7 (6%) | 6 (7%) | 1 (4%) | 0.499 |
| **Tobacco Exposure History** | Never Smoker | 77 (69%) | 63 (75%) | 14 (50%) | 0.034 |
|  | Former Smoker | 20 (18%) | 14 (17%) | 6 (21%) |  |
|  | Current Smoker | 7 (6%) | 3 (4%) | 4 (14%) |  |
|  | Unknown | 8 (7%) | 4 (5%) | 4 (14%) |  |

All data reported as n(%) with exception of age which is reported as Median (IQR). reported p-value is from Chi Squared test with exception of age where a two-sided t-test was used. PMHx: Past Medical History; COPD: Chronic Obstructive Pulmonary Disease; CAD: Coronary Artery Disease; CKD: Chronic Kidney Disease.

### Rate of Admissions

As of April 17, the day after the last COVID diagnosis for this cohort, there were 11,391 confirmed COVID-19 cases reported in Los Angeles County. During this period, there were 113 unique COVID related admissions in our hospital system at a rate of 1-8 patients per day. The peak number of new COVID-related admissions was 8 on April 3rd (Supplemental Figure 1).

### Inflammatory Markers on Admission

Our cohort had a notable elevation of pro-inflammatory markers on presentation without an elevated white blood count (median 6.25 x 10^3^/μL IQR 4.75-8.55 x 10^3^/μL). Median interleukin-6 level was 9 pg/ml on admission (upper limit of normal for assay <5 pg/mL) (IQR 2-23 pg/ml), median D-dimer was 1140 ng/ml (IQR 677-2073 ng/ml), and median ferritin level was 696 ng/ml (IQR= 357-1616 ng/ml). The median absolute lymphocyte count was 0.9 x 10^3^/μL (IQR 0.58-1.18 x 10^3^/ μL), median CRP was 7.8 mg/dl (IQR 3.2-12.65 mg/dl), median LDH was 324 U/L (IQR 239-423), and median procalcitonin was 0.12 ug/L (IQR 0-0.385 ug/L) (Table 3).

### Patient Outcomes

Of the 112 patients evaluated, there were 47 ICU admissions, 28 intubations and 7 deaths. Of the 112 patients, all had completed disposition at the time of this report. The rate of in-hospital 60-day mortality for our total cohort was 6% (7/112) with an ICU mortality of 12.8% (6/47), and a mortality of 18% (5/28) for those who required endotracheal intubation. Two patients received CPR, neither of whom achieved return of spontaneous circulation. Of the 7 patients who died, 2 had a do not resuscitate (DNR) order documented in the chart at the time of admission. Median ICU length of stay (LOS) was 7 days (IQR 3-15 days).

### Course of Respiratory Failure

We evaluated individual patients’ trajectories of respiratory failure. High-flow nasal cannula (HFNC) was required for 11 patients, of whom 45% progressed to intubation. Conversely, of those who were intubated, only 18% were previously on HFNC. Baseline factors associated with need for intubation were obesity (OR 5.82, p=0.004), former smoker status (OR 8.06, p=0.02), and current smoking status (OR 10.33, p=0.02), as per multivariable logistic regression analysis adjusting for age, sex, and high prevalence comorbidities (Table 2, Figure 2). Additionally, the proinflammatory markers IL-6, ferritin, LDH and CRP, drawn on admission or on day of COVID-19 diagnosis for patients already hospitalized at the time of positive PCR, were associated with higher odds of intubation by univariate analysis (Table 3). The absolute lymphocyte count was also associated with need for mechanical ventilation (OR of 0.33, CI 0.11-0.96).

**Table 2.**
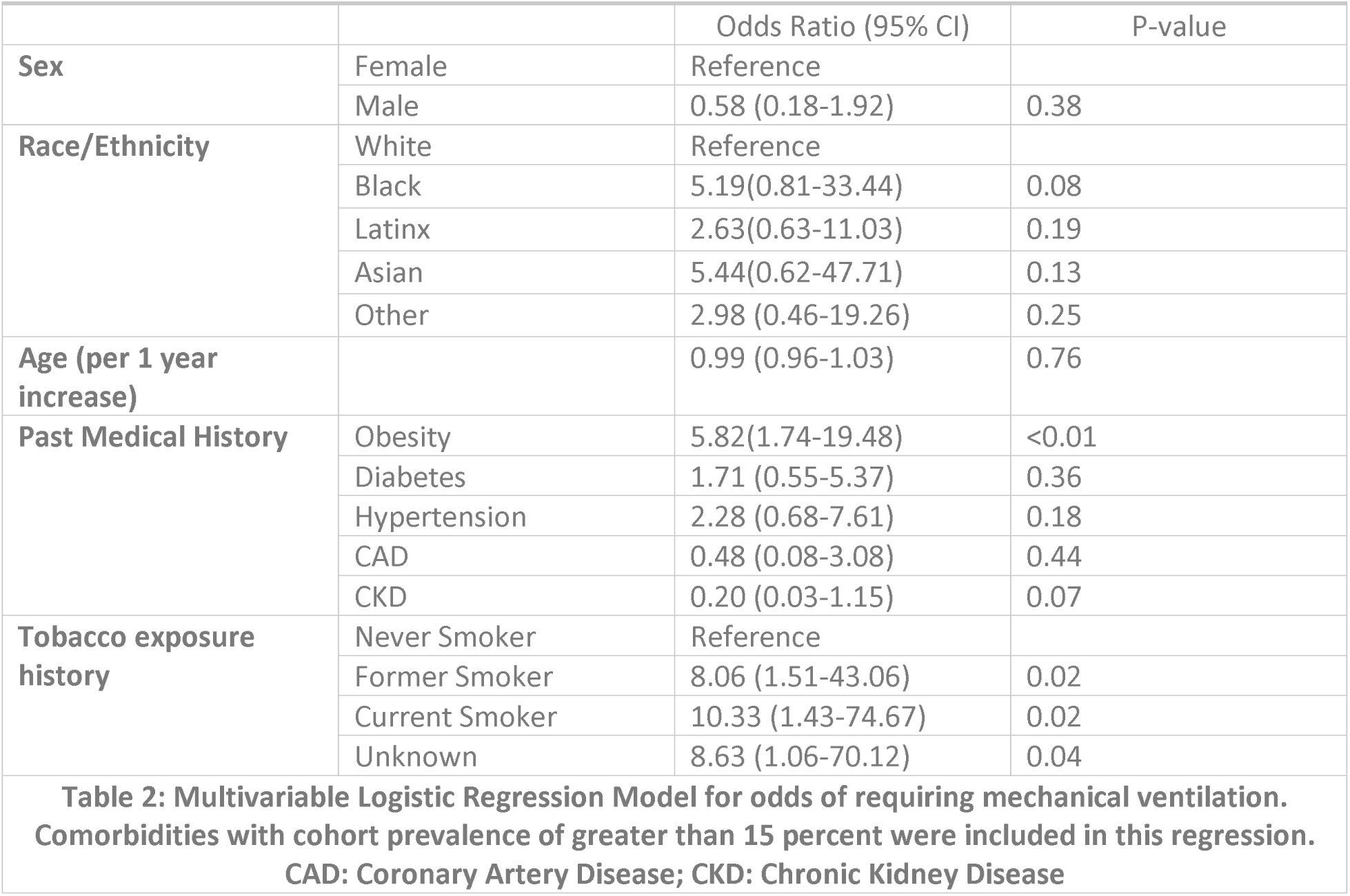
Multivariable Logistic Regression for odds of requiring mechanical ventilation.

**Table 3.** Univariate logistic regression model for requiring mechanical ventilation.

|  |  |  |  |  |
| --- | --- | --- | --- | --- |
|  | N | Median (IQR) | Odds Ratio (95% CI) | P-value |
| Absolute Lymphocyte Count (10<sup>3</sup>/uL ) | 111 | 6.25 (4.8-8.5) | 0.33 (0.11-0.96) | 0.042 |
| LDH (U/L)* | 102 | 324 (239-423) | 1.57 (1.13-2.17) | 0.006 |
| Ferritin (ng/ml)* | 98 | 696 (357-1616) | 1.05 (1.005-1.11) | 0.032 |
| C-Reactive Protein (mg/dl) | 100 | 7.8 (3.2-12.65) | 1.13 (1.06-1.21) | <0.001 |
| D-Dimer (ng/ml)* | 99 | 1140 (677-2073) | 1.02 (0.99-1.04) | 0.053 |
| Procalcitonin (µg/L) | 108 | 0.12 (0-0.385) | 1.27 (1.02-1.57) | 0.030 |
| Interleukin-6 (pg/ml)** | 91 | 9 (2-23) | 1.17 (1.03-1.33) | 0.015 |
| *per 100-unit change of inflammatory marker. **per 10-unit change of inflammatory marker. |  |  |  |  |

Out of the 28 patients intubated, 24 patients were diagnosed with ARDS (21% of our total cohort), and 21 patients had a P/F ratio < 150 (19% of our total cohort). An additional three patients had bilateral opacities and required HFNC. The ICU LOS was significantly greater amongst patients with ARDS. Those with ARDS had an ICU LOS of 15 days (IQR 9-24 days) while those without ARDS had an ICU LOS of 5 days (IQR 2-5 days; difference with p< 0.01).

### Pharmacologic Therapies

During the period of data collection, there was no definitive evidence supporting or discrediting the use of selected anti-inflammatory or anti-viral medications for the treatment of COVID-19. In our cohort, 54% of patients were either enrolled in a placebo-controlled trial or received open label use of one or more of the following therapies: anti-IL-6 therapy (sarilumab or tocilizumab), CCR5 antagonist (leronlimab), hydroxychloroquine, or remdesivir (Supplemental Tables 1 and 2). Mortality between the patients who received at least one of these interventions/studies was statistically similar to those who were not subject to open label interventions or trial enrollment, with 2 deaths in the group of 60 patients enrolled in a trial or receiving open label therapy versus 5 deaths out of the 52 who did not (p=0.171 by chi squared, unadjusted analysis). The correlation of any individual therapy with mortality could not be determined due to the overall low mortality rate of our cohort.

Treatment guidance at our institution recommended the use of pharmacologic VTE prophylaxis on all patients unless contraindicated. Therapeutic anticoagulation for VTE was recommended only in the case of confirmed or highly suspected VTE. In our cohort, 26/112 received therapeutic dose anticoagulation; 25 of these patients (96%) had a known indication for therapeutic anticoagulation, with only 1 patient receiving anticoagulation empirically. Indications for therapeutic anticoagulation included chronic conditions (atrial fibrillation, bioprosthetic valve or prior VTE, totaling 7 patients), new atrial fibrillation (6 patients), and need for extracorporeal support (1 patient). Eleven patients had suspected or confirmed acute thromboembolic events during their hospitalization, including deep venous thrombosis or pulmonary embolus (6 patients), frequent clotting of dialysis catheters (2 patients), acute coronary syndrome (1 patients), ischemic digit (1 patient), or left ventricular thrombus (1 patient) (Supplemental Table 3). Of the 6 patients (5.4%) who developed radiographically-confirmed VTE, 2 were diagnosed at the time of admission. Of the 4 who later developed VTE, all were on VTE prophylaxis at the time of diagnosis. Three of the patients with radiographically-confirmed VTE had ARDS. None of the patients with new VTE died during their hospitalization. Mortality was not significantly different amongst those receiving and not receiving therapeutic anticoagulation (4% v. 7%, χ^2^ p=0.563).

While there has been debate about the use of steroids in the treatment of COVID-19, our institutional treatment guidance did not recommend routine steroids for COVID-19 related ARDS in the absence of another indication during this time period. From our cohort, 11/112 received steroid doses that exceeded 20 mg of prednisone equivalents daily. Of those, 10/11 had an indication based on past medical history (e.g., chronic steroid use), new adrenal insufficiency, or refractory shock.

### ARDS Management

In patients who developed ARDS, we examined the rate of adherence to lung protective ventilation and the frequency of adjunctive therapy use. Patients with ARDS had a median tidal volume of 6.1 cc/kg PBW on day 1. The pooled median tidal volumes per PBW in days 2-7 varied from 5.9 to 6.1 (Figure 1). Outliers with larger tidal volumes tended to be those patients placed on pressure support. Adherence to LPV (<8cc/kg PBW) was 90.2% for ventilator days 1-7. We also evaluated the use of PEEP in our cohort. For patients with mild to moderate ARDS (P/F > 150, n=3), median PEEP was between 5-10 cmH_2_O depending on the day for the first 7 days of mechanical ventilation. For moderate to severe ARDS (P/F ratio < 150, n=21), median daily PEEP was 10-12 cmH_2_O for the first 7 days of mechanical ventilation.

**Figure 1:**
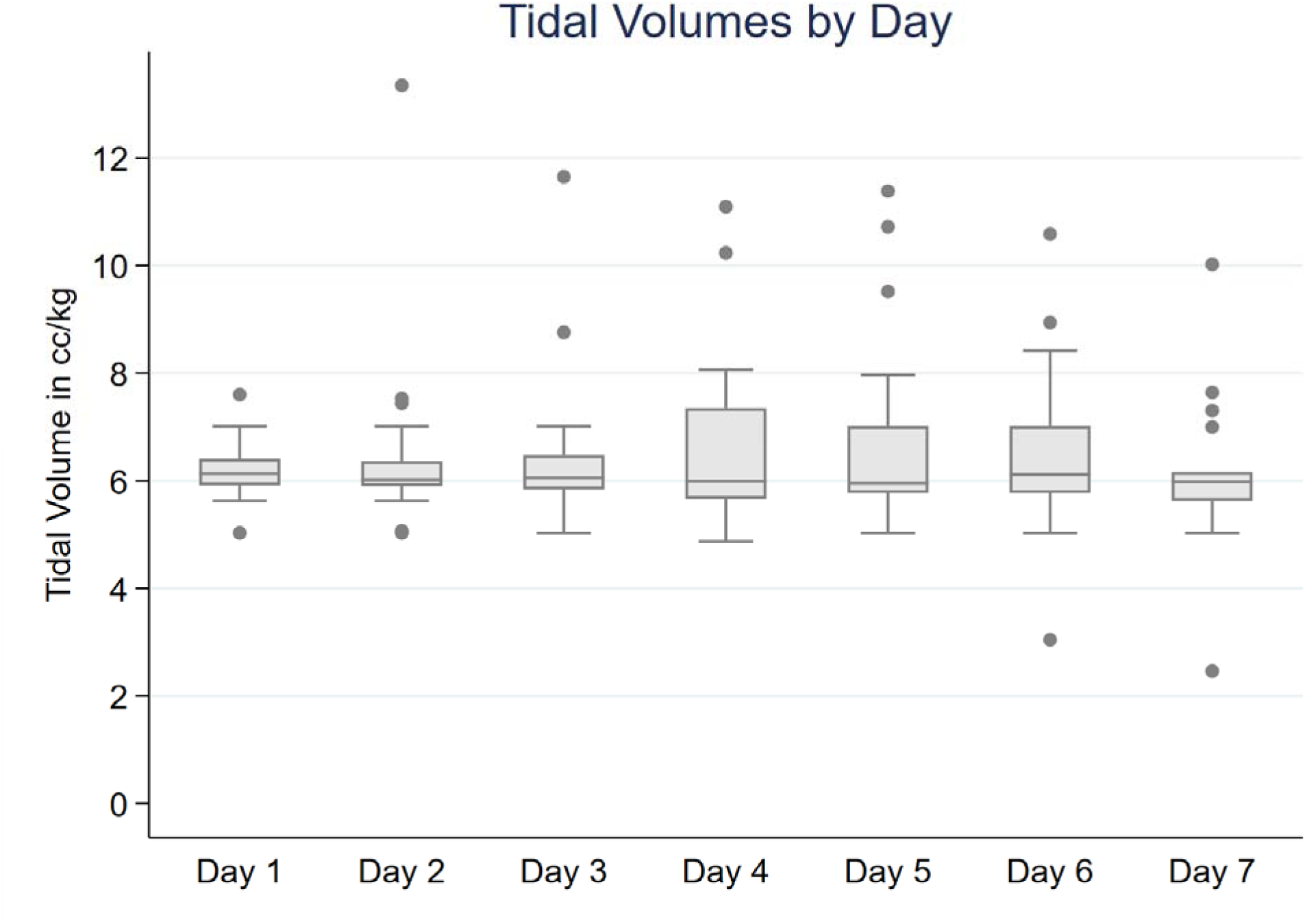
Box and whisker plot of pooled cohort mechanical ventilation tidal volume for first 7 days postintubation. Tidal volume is displayed as cc/kg of predicted body weight. Box represents IQR and whiskers represent minimum and maximum, with outliers represented as dots.

**Figure 2:**
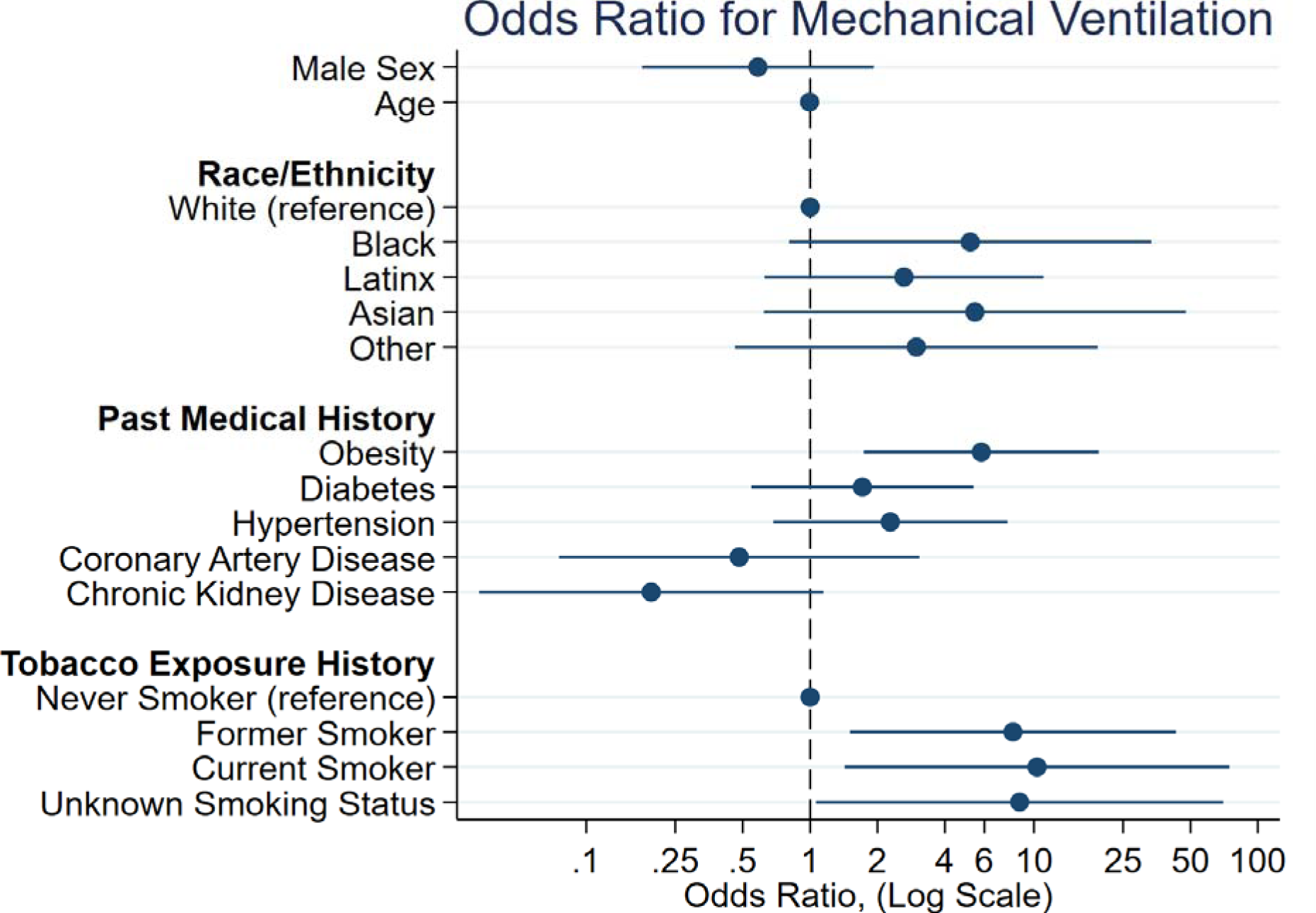
Forest plot of multivariable logistic regression analysis adjusting for age, sex, and comorbidities with cohort prevalence of >15%. The x-axis is depicted on a log scale.

Out of 21 patients with ARDS and P/F ratio < 150, 12 patients (57%) underwent prone positioning at least once. Among those with available data (n= 9), the P/F ratio improved by an average of 63 points (SD 84) after the first proning event. In addition, out of the 28 intubated patients, 14/28 (50%) received neuromuscular blockade outside of rapid sequence intubation. Only a minority (2/28) of intubated patients received inhaled nitric oxide at any point, and only one patient received veno-venous extracorporeal membrane oxygenation (VV-ECMO).

## Discussion

In this retrospective, observational study, we report lower hospital mortality in both critically and non-critically ill patients at our center in comparison with prior published cohorts. Patient-level characteristics associated with need for invasive mechanical ventilation included obesity, past and present smoking history, and elevation of pro-inflammatory markers. Further understanding of the factors associated with clinical deterioration may help identify therapeutic targets for early intervention.

To our knowledge, this is the first study identifying obesity and smoking history as risk factors for the need for invasive mechanical ventilation in COVID-19. While obesity has been associated with increased risk for a positive COVID-19 test,^11^ and an association between obesity and mortality in COVID-19 has been speculated,^12,13^ its association with outcomes in COVID-19 has not been directly evaluated. On the other hand, the impact of smoking in COVID-19 disease severity may be explained by the observation that smokers have upregulation of ACE2 receptors on lung biopsy.^14,15^ Indeed, a large observational cohort from Wuhan reported a higher rate of smoking among patients with more severe forms of COVID-19.^16^ In addition, a smaller study revealed a higher adjusted risk for disease progression in patients with a history of smoking compared to never smokers.^17^ Metanalyses evaluating the relationship between smoking and COVID-19 disease severity have had mixed results. A smaller evaluation of five published cohorts from China did not find a statistically significant association between active smoking and disease severity; however the analysis was unadjusted.^18^ A larger metanalysis found that current or former smoking was associated higher unadjusted risk for the composite outcome of ICU admission, intubation or death in COVID-19.^19^ None of the included studies examined adjusted risk for IMV.^19,20^

Overall mortality in our patient population was low. While the factors associated with mortality remain unclear, this finding is encouraging. This cohort shared some similarities to others, including a majority of male patients (66%),^16,21-24^ a median age of 61,^21,23,24^ and an admission rate of 56% nonwhite minorities.^24^ We also report comparable rates of invasive mechanical ventilation to other cohorts.^21,24^ However, the role of baseline severity of illness was not fully assessed, and has not been extensively detailed in prior studies. As such, it is possible that our patient population was less ill than others.

In our ventilated patients, the in-hospital mortality rate was 18%, which was similar to the rate seen in a recently published study, but lower than that seen in earlier cohorts.^23-25^ The vast majority of mechanically ventilated patients in our cohort had ARDS, suggesting that optimizing ARDS care may improve the outcomes of these patients. ARDS is traditionally underdiagnosed worldwide, and prior studies demonstrate the underutilization of therapies known to reduce mortality in this syndrome.^26,27^

Both lung protective ventilation and prone positioning have been associated with improved outcomes in ARDS and are recommended by multiple professional society guidelines.^6,7,28,29^ Our adherence to lung protective ventilation strategies (90.2% for ventilator days 1-7) was higher than that reported in previous large multicenter ARDS cohorts.^27,30^ Prone positioning was also used almost 60% of the time compared with 8-11% in prior reports.^26,27^ While we were unable to definitively correlate any individual therapeutic measure with mortality given the small size of our cohort, adherence to established ARDS best practices may have played a role. It is also likely that the lower per capita incidence of COVID-19 in our state^31^ and the lack of strain in our system enabled adherence to these labor-intensive interventions for ARDS. Indeed, as of the time of this writing, there was no change to usual staffing ratios, or need for non-ICU clinicians to take care of ICU level patients. Although this finding warrants further investigation, it may suggest that efforts to mitigate the spread of COVID-19 as a means to avoid straining healthcare systems may indeed be an effective strategy for controlling mortality from this disease.

Regarding features that may be specific to COVID-19, there have been several reports of high rates of coagulopathy and VTE in these patients.^32-34^ Post-mortem studies revealed a high rate of microthrombi in the pulmonary vasculature of COVID-19 patients ^35^ Microvascular damage and thrombi formation are in fact well documented in ARDS.^36,37^ We found a low rate of VTE (5%) in our patient population. While VTE was not a focus of our study, some mitigating factors associated with its seemingly low incidence in our cohort may include a high use of VTE prophylaxis, as well as recommendations by our pandemic response team for daily awakenings, light sedation, and regular consultation with physical and occupational therapy.

Recent reports have also suggested that COVID-19 produces an excessive inflammatory response by the host. Indeed, IL-6 and serum ferritin, traditionally elevated in proinflammatory conditions, have been associated with higher mortality in patients infected with COVID-19.^16^ In this cohort, ferritin and IL-6 also correlated with increased rates of endotracheal intubation, but no definitive correlation with mortality could be made. Given the concern that excessive inflammatory response may play a role in mortality, multiple clinical trials have been initiated to inhibit a number of targets including interleukin 1, interleukin 6, and granulocyte-macrophage-colony stimulating factor. While a large portion of our patients were enrolled in clinical trials, the impact of these agents remains to be seen.

The strengths of this study included manual extraction of all data points, which allowed for higher accuracy and granularity of findings. In addition, the in-hospital mortality was known for all of our patients. Study weaknesses included the single center nature and small sample size, which precluded more robust evaluation of risk factors associated with mortality. In addition, short follow up times precluded the assessment of disease sequela in our cohort. We also did not collect information on social determinants of health such as education level, insurance status or economic factors, all of which could have influenced observed outcomes. Finally, given its retrospective nature, we also cannot ascertain that any of our interventions directly affected patient outcomes. Nevertheless, our data identified potential risk factors for disease progression.

## Conclusion

Early experience with COVID-19 at UCLA has revealed a lower mortality than previously reported. Baseline factors associated with increased odds for the need for mechanical ventilation include obesity and smoking history; further research is needed to confirm these findings in larger, multicenter cohorts. Finally, the incidence of ARDS among intubated patients with COVID-19 is high, suggesting that optimizing ARDS care may improve the outcomes of these patients.

## Data Availability

The dataset used for this manuscript will be readily available upon request to the corresponding author.

---

**Supplemental Table 1.** Experimental Therapies Administered to Cohort Patients.

|  |  |
| --- | --- |
| Medication | Number of patients |
| No Medication | 52 |
| **Individual Therapies** |  |
| Anti-IL-6 | 3 |
| Remdesivir | 6 |
| Leronlimab | 6 |
| Hydroxychloroquine | 15 |
| **Combination Therapies** |  |
| Anti-IL-6 and Remdesivir | 1 |
| Anti-IL-6 and Leronlimab | 3 |
| Anti-IL-6 and Hydroxychloroquine | 14 |
| Remdesivir and Leronlimab | 1 |
| Remdesivir and Hydroxychloroquine | 2 |
| Leronlimab and Hydroxychloroquine | 2 |
| Anti-IL-6 and Leronlimab and Hydroxychloroquine | 1 |
| Anti-IL-6 and Remdesivir and Hydroxychloroquine | 5 |
| Anti-IL-6 and Remdesivir and Leronlimab and Hydroxychloroquine | 1 |
| *\*Anti-IL 6 therapies include tocilizumab and sarilumab. Patients in these groups received only one type.* |  |

**Supplemental Table 2:** Interventions administered open label or via trial enrollment. Those enrolled in clinical trial received either the listed medication or placebo in a blinded fashion.

|  |  |  |  |
| --- | --- | --- | --- |
| Medication | Total | % in Trial | % Open Label |
| Hydroxychloroquine | 40 | 0% | 100% |
| Leronlimab | 14 | 7% | 93% |
| Remdesivir (or placebo) | 16 | 88% | 22% |
| Sarilumab (or placebo) | 9 | 100% | 0% |
| Tocilizumab | 19 | 21% | 79% |

**Supplemental Table 3:** Reported Indication for Therapeutic Dose Anticoagulation.

|  |  |
| --- | --- |
| Anticoagulation Indication | N |
| Acute Coronary Syndrome | 1 |
| Atrial Fibrillation | 6 |
| ECMO | 1 |
| Empiric for <24 hours | 1 |
| Frequent dialysis line clotting | 2 |
| Ischemic toe | 1 |
| LV Thrombus | 1 |
| New in-hospital VTE | 6 |
| Past Medical History (includes prior VTE) | 7 |

**Supplemental Figure 1:**
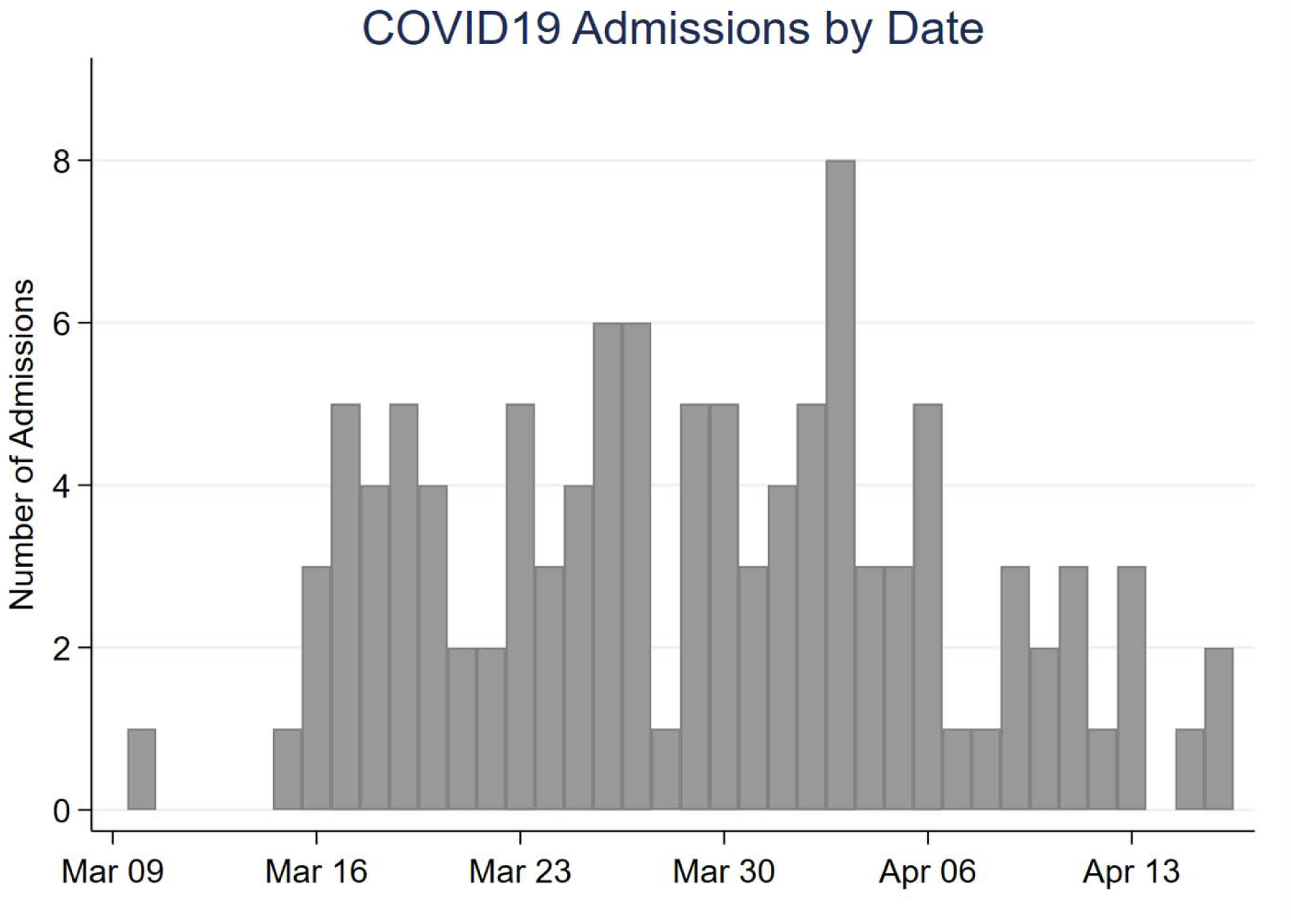
Combined COVID-19 admissions by date for both Ronald Reagan UCLA and Santa Monica UCLA. Of note, one admission from December and one from February, both of whom contracted COVID-19 during hospitalization were omitted from this histogram for ease of interpretation.

